# Calibration Drift Under Cross-Institutional Deployment: An External Validation Framework for ICU Mortality Prediction Across MIMIC-IV and eICU

**DOI:** 10.64898/2026.05.03.26352335

**Authors:** Krutarth Patel, Phanindra Beedala

## Abstract

**Background:** Machine learning models for intensive care unit (ICU) mortality prediction achieve strong internal discrimination yet rarely undergo external validation with calibration assessment — a gap undermining clinical deployment. Calibration, the agreement between predicted probabilities and observed event rates, is prerequisite for threshold-based decisions yet remains underreported.

**Methods:** We conducted a retrospective cohort study using MIMIC-IV (v2.2; n = 52,028 ICU stays) for model development and eICU (n = 114,060) for independent external validation. Logistic regression, random forest, and gradient boosting (XGBoost) were evaluated on first-24-hour clinical variables. Discrimination was assessed via receiver operating characteristic area (AUROC) and precision-recall area (AUPRC); calibration via slope, intercept, and expected calibration error (ECE). Post-hoc logistic recalibration was applied externally. Clinical utility was evaluated by decision curve analysis benchmarked against Acute Physiology and Chronic Health Evaluation (APACHE) scores. Subgroup analyses examined sex and race/ethnicity; SHapley Additive exPlanations (SHAP) assessed feature importance. Uncertainty was estimated via bootstrap resampling; the study adheres to TRIPOD guidelines.

**Results:** The recalibrated XGBoost model achieved internal AUROC 0.847 (95% CI: 0.832–0.860) and external AUROC 0.819 (95% CI: 0.815–0.823). Internal calibration was near-ideal (slope 0.982; intercept 0.001), whereas external validation revealed systematic risk overestimation (intercept −0.678) attributable to prevalence-driven label shift. An intercept-only adjustment reduced ECE by 26%. The model outperformed APACHE (AUROC 0.817 vs. 0.795; p < 0.001).

**Conclusions:** ICU mortality models exhibit transportable discrimination but clinically significant calibration drift under cross-institutional deployment. Calibration evaluation and targeted recalibration should be mandatory in any clinical machine learning validation framework.

## 1. INTRODUCTION

Machine learning models for intensive care unit (ICU) mortality prediction routinely achieve strong internal discrimination yet fail to meet calibration standards when transferred across institutions — a gap with direct and underappreciated consequences for clinical deployment [1, 2]. Reliable risk stratification can guide triage, inform resource allocation, and support high-stakes bedside decisions [3, 4], but a model that correctly ranks patients by relative risk may simultaneously overestimate or underestimate absolute mortality probability by a clinically significant margin [5]. This dissociation between preserved discrimination and degraded calibration is the central deployment hazard this work addresses.

The widespread adoption of electronic health records has enabled data-driven outcome prediction [6], and machine learning techniques have demonstrated the ability to model complex nonlinear relationships across physiological, laboratory, and demographic variables [7, 8]. Large critical care databases — including MIMIC-IV and the eICU Collaborative Research Database — have facilitated the development and benchmarking of ICU mortality models [9, 10], with logistic regression, random forest, and gradient boosting approaches consistently reporting strong discriminative performance [11, 12]. Yet single-dataset performance does not guarantee reliable generalization: differences in patient populations, clinical workflows, and data collection practices can substantially alter model behavior in external cohorts [13], and systematic reviews have documented that external validation remains inconsistently reported across the clinical prediction model literature [14] — making rigorous cross-institutional evaluation essential for assessing deployment readiness [15].

The field faces additional concerns regarding reproducibility and methodological rigor [16]. Variability in cohort definitions and preprocessing pipelines produces non-comparable results, and temporal data leakage can generate overly optimistic performance estimates [17]. Most critically, while AUROC is widely reported, calibration — the agreement between predicted probabilities and observed event rates — remains systematically under-evaluated despite its direct importance for threshold-based clinical decisions [18, 19]. Emerging evidence confirms that distributional shifts between institutions disproportionately affect absolute probability estimates while leaving discriminative ranking relatively intact [18], a dissociation with serious implications for any deployment context where absolute risk estimates drive clinical action.

To address these gaps, this study presents a reproducible, calibration-aware benchmarking framework for ICU hospital mortality prediction, operationalized across two large, independent, publicly available critical care databases. The framework spans cohort construction, feature extraction, preprocessing, model development, calibration-aware evaluation, post-hoc recalibration, and demographic fairness auditing — with all code, cohort definitions, and model artifacts publicly available as an immediately adoptable evaluation template for clinical informatics researchers.

## 2. METHODS

### 2.1 Study Design and Data Sources

We conducted a retrospective cohort study using two publicly available, de-identified critical care databases. MIMIC-IV (version 2.2) [9] comprises longitudinal EHR data from a single quaternary academic medical center (Beth Israel Deaconess Medical Center, Boston, MA, USA; 2008–2022) and served as the model development and internal validation source. The eICU Collaborative Research Database [10] aggregates ICU data from 335 units across 208 U.S. hospitals (2014–2015) and constituted the independent external validation cohort. All procedures adhered to established standards for clinical prediction model development and external validation [13].

The primary outcome was in-hospital mortality; secondary outcomes were ICU mortality and prolonged ICU length of stay (≥7 days). Eligibility criteria were applied identically across both datasets: adult patients (aged ≥18 years) with a unique first ICU stay per hospitalization. Exclusion criteria were: ICU stays <4 hours, missing outcome data, and age <18 years. As all data were fully de-identified and publicly available under approved data-use agreements, IRB review was not required.

Baseline cohort characteristics are reported in Table 1. Standardised mean differences (SMDs) were computed to characterise distributional similarity between the development and external validation cohorts: |μ□ − μ□| / pooled SD for continuous normally distributed variables, the proportion-based formula for binary variables, and raw individual-level data before aggregation for non-normally distributed variables reported as median [IQR].

**Table 1.** Baseline demographic and clinical characteristics of the MIMIC-IV and eICU study cohorts.

| Variable | MIMIC-IV (n = 52,028) | eICU (n = 114,060) | SMD |
| --- | --- | --- | --- |
| **Demographics** |  |  |  |
| Age, mean (SD), years | 63.61 (16.56) | 63.69 (16.77) | 0.005 |
| Female, % | 43.69 | 46.01 | 0.046 |
| **Vital signs** |  |  |  |
| Heart rate, mean (SD), bpm | 85.00 (15.76) | 84.99 (16.38) | <0.01 |
| MAP, mean (SD), mmHg | 77.61 (12.20) | 81.72 (13.48) | 0.322 |
| Respiratory rate, mean (SD) | 19.15 (3.76) | 19.66 (4.74) | 0.12 |
| SpO<sub>2</sub>, mean (SD), % | 96.95 (1.99) | 96.85 (2.21) | 0.048 |
| **Laboratory values** |  |  |  |
| BUN, median [IQR], mg/dL | 20.00 [13.00–32.00] | 20.00 [13.00–34.00] | 0.037 |
| Creatinine, median [IQR], mg/dL | 1.00 [0.70–1.50] | 1.03 [0.76–1.63] | 0.074 |
| Lactate, median [IQR], mmol/L | 2.10 [1.40–3.10] | 1.80 [1.20–3.10] | 0.183 |
| Hemoglobin, median [IQR],<br>g/dL | 10.00 [8.50–11.70] | 10.70 [9.00–12.40] | 0.157 |
| Platelets, median [IQR],<br>×10<sup>3</sup>/μL | 177 [125–240] | 185 [135–243] | 0.065 |
| **Clinical indicators and outcomes** |  |  |  |
| Urine output 24 h, median<br>[IQR], mL | 2160 [1300–3250] | 1335 [730–2120] | 0.431 |
| Hospital mortality, % | 10.5 | 8.69 | 0.06 |
| ICU mortality, % | 6.71 | 5.55 | 0.047 |
| Prolonged ICU LOS ≥7 days, % | 13.19 | 10.91 | 0.072 |

Values are mean (SD), median [IQR], or percentage as appropriate. SMD = standardised mean difference; SMD <0.10 indicates negligible distributional difference. Race/ethnicity was not available in harmonised form for MIMIC-IV and is reported for eICU only in Section 3.5. MAP = mean arterial pressure; BUN = blood urea nitrogen; LOS = length of stay. The large urine output SMD (0.431) likely reflects institutional variation in fluid protocols and documentation practices rather than a fundamental cohort difference.

### 2.2 Feature Extraction and Preprocessing

Predictor variables were restricted to routinely collected clinical data from the first 24 hours of ICU admission to reflect realistic deployment constraints and prevent temporal data leakage [15, 16, 17]. Features spanned four domains: demographics, vital signs, laboratory measurements, and clinical treatment indicators including fluid balance. Thirty-six candidate features were initially extracted; after screening, the final modeling set comprised 38 predictors inclusive of binary missingness indicators (added for variables with >10% missing values). Continuous variables were summarized using minimum, maximum, or mean within the 24-hour window (detailed in Supplementary Table S1).

All preprocessing parameters — imputation values, encoding mappings, and scaling transformations — were derived exclusively from the MIMIC-IV training partition and applied without modification to all validation and external cohorts [13]. Features with >45% missingness were excluded; remaining missing values were imputed using training-set medians [13]. Categorical variables were one-hot encoded; min–max normalization was applied to logistic regression only.

### 2.3 Model Development

MIMIC-IV was partitioned into training (70%), internal validation (15%), and held-out test (15%) sets using stratified random sampling (random seed = 42). The eICU dataset was withheld entirely for external validation. Three model classes were evaluated: logistic regression, random forest [20], and gradient boosting (XGBoost). Class imbalance (∼10% mortality rate) was addressed through class-weighted loss functions.

Given that calibration is a critical and under-evaluated dimension of clinical model validity [18, 19], model selection was based on a study-specific composite criterion: Score = 0.40 × AUROC + 0.25 × AUPRC − 0.20 × Brier − 0.10 × ECE − 0.03 × |slope − 1| − 0.02 × |intercept|, where ECE denotes expected calibration error [18, 19]. Higher scores indicate better overall performance, with weights selected to prioritize discrimination while explicitly penalizing miscalibration and probabilistic error. Hyperparameters were tuned via five-fold stratified cross-validation; the binary threshold was set using the Youden index. The XGBoost model with post hoc logistic recalibration achieved the highest composite validation score and was designated the primary model.

### 2.4 Validation and Evaluation

### Internal validation

was performed on the held-out MIMIC-IV test set. Metrics comprised: discrimination (AUROC, AUPRC [21, 22]); overall probabilistic accuracy (Brier score [23]); calibration (slope, intercept, ECE [18, 19]); and binary classification performance (sensitivity, specificity, PPV, NPV, F1). Bootstrap confidence intervals were estimated using 500 iterations for primary model-performance analyses, 300 iterations for subgroup analyses, and 1,000 iterations for paired benchmark comparisons. Iteration counts were selected to balance computational efficiency with estimation stability across analyses of varying complexity.

### External validation

was conducted on the full eICU cohort without retraining or parameter updates, applying all preprocessing exactly as defined on MIMIC-IV training data [15]. Post hoc Platt scaling was applied to external predictions using internal validation outputs [24, 25]; a label-shift intercept-only adjustment was evaluated as a sensitivity analysis. Calibration was assessed graphically (loess-smoothed calibration curves) and quantitatively. Clinical utility was evaluated using decision curve analysis (DCA), quantifying net benefit across clinically plausible threshold probabilities relative to treat-all and treat-none strategies [26, 27], with additional benchmark comparison against APACHE scores in a matched eICU subset.

### 2.5 Subgroup, Sensitivity, and Interpretability Analyses

Subgroup analyses were performed across sex and race/ethnicity groups available in eICU. AUROC and calibration metrics were computed per subgroup with bootstrap confidence intervals; performance disparities were quantified as absolute between-group differences following established algorithmic fairness frameworks [28, 29]. Five pre-specified sensitivity analyses assessed robustness to: single-stay-per-patient restriction; restriction to ICU stays ≥48 hours; exclusion of laboratory variables; exclusion of arterial blood gas features; and exclusion of race/ethnicity variables.

Model interpretability was assessed using SHAP values (TreeExplainer) [30] and permutation importance, with rank concordance evaluated by Spearman correlation.

### 2.6 Reproducibility and Reporting

All analyses were implemented in Python (random seed = 42). The complete pipeline is publicly available at https://github.com/Krutarth007/icu-mortality-prediction-ml. This study adheres to TRIPOD reporting guidelines [31] and incorporates PROBAST risk-of-bias assessment [32], consistent with standards for rigorous clinical machine learning research [16].

## 3. RESULTS

### 3.1 Cohort Characteristics

After applying eligibility criteria, the development cohort comprised 52,028 adult ICU stays from MIMIC-IV (training n = 36,328; validation n = 7,933; held-out test n = 7,767) and the external validation cohort comprised 114,060 ICU stays from eICU. In-hospital mortality was 10.5% in MIMIC-IV and 8.7% in eICU. Baseline characteristics are presented in **Table 1**. Standardised mean differences (SMDs) were <0.10 for most variables, indicating broadly comparable distributions; the largest shifts were observed for 24-hour urine output (median 2,160 vs. 1,335 mL; SMD = 0.431) and mean arterial pressure (77.6 vs. 81.7 mmHg; SMD = 0.322), likely reflecting institutional differences in fluid management protocols and documentation practices.

### 3.2 Model Performance

Internal and external performance metrics are summarised in **Table 2**. On the held-out MIMIC-IV test set, the primary model (XGBoost + logistic recalibration) achieved AUROC 0.847 (95% CI 0.832–0.860), AUPRC 0.441 (95% CI 0.402–0.475), and Brier score 0.075 (95% CI 0.071–0.079). In external validation on eICU, the model maintained strong discrimination (AUROC 0.819, 95% CI 0.815–0.823; AUPRC 0.355; Brier 0.072), with an absolute AUROC reduction of 0.028 — consistent with expected attenuation under cross-institutional distributional shift. ROC curves for all models on the internal test set are shown in **Figure 2**.

**Figure 1.**
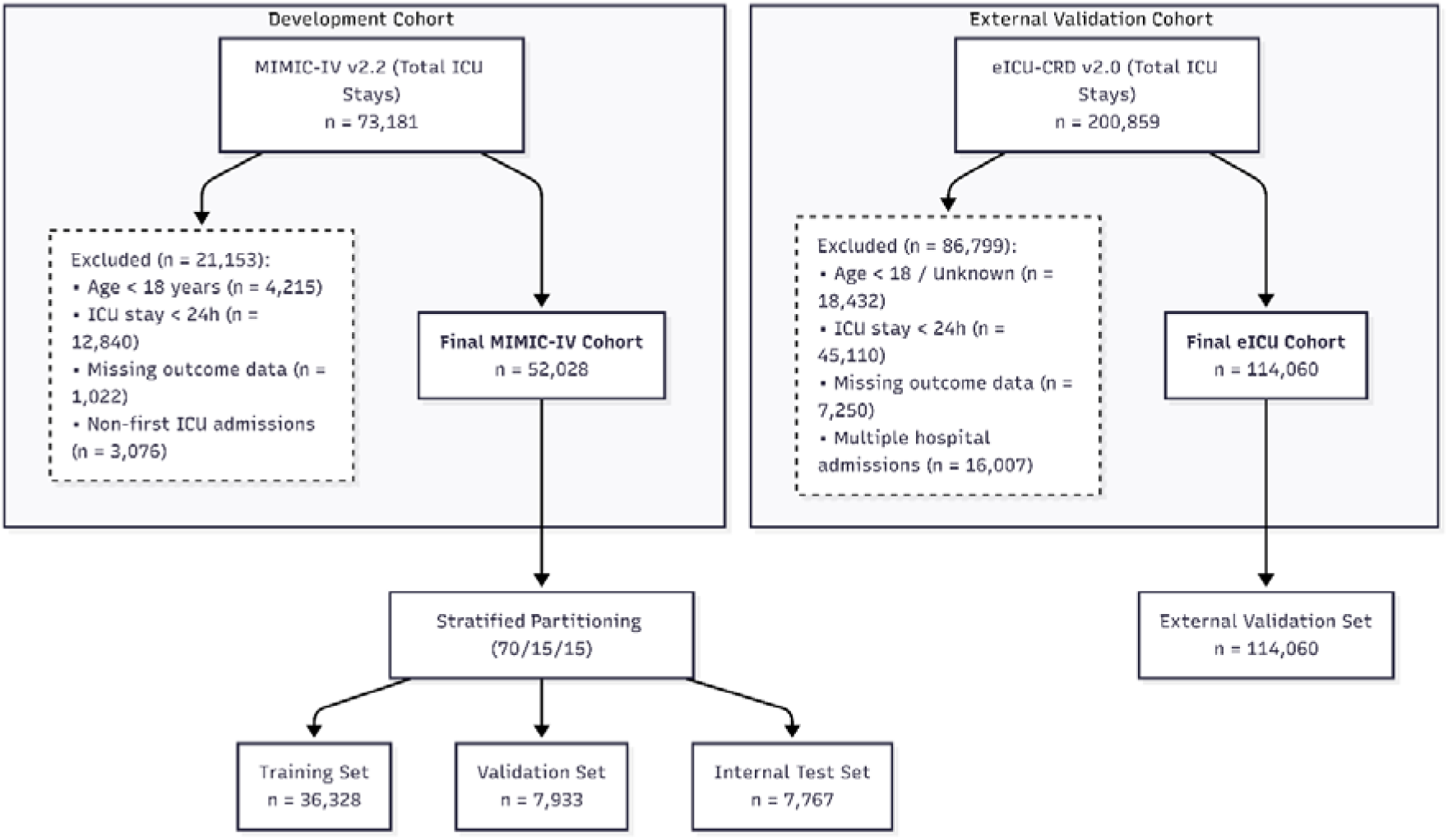
Cohort selection and data partitioning for model development (MIMIC-IV) and external validation (eICU)

**Figure 2.**
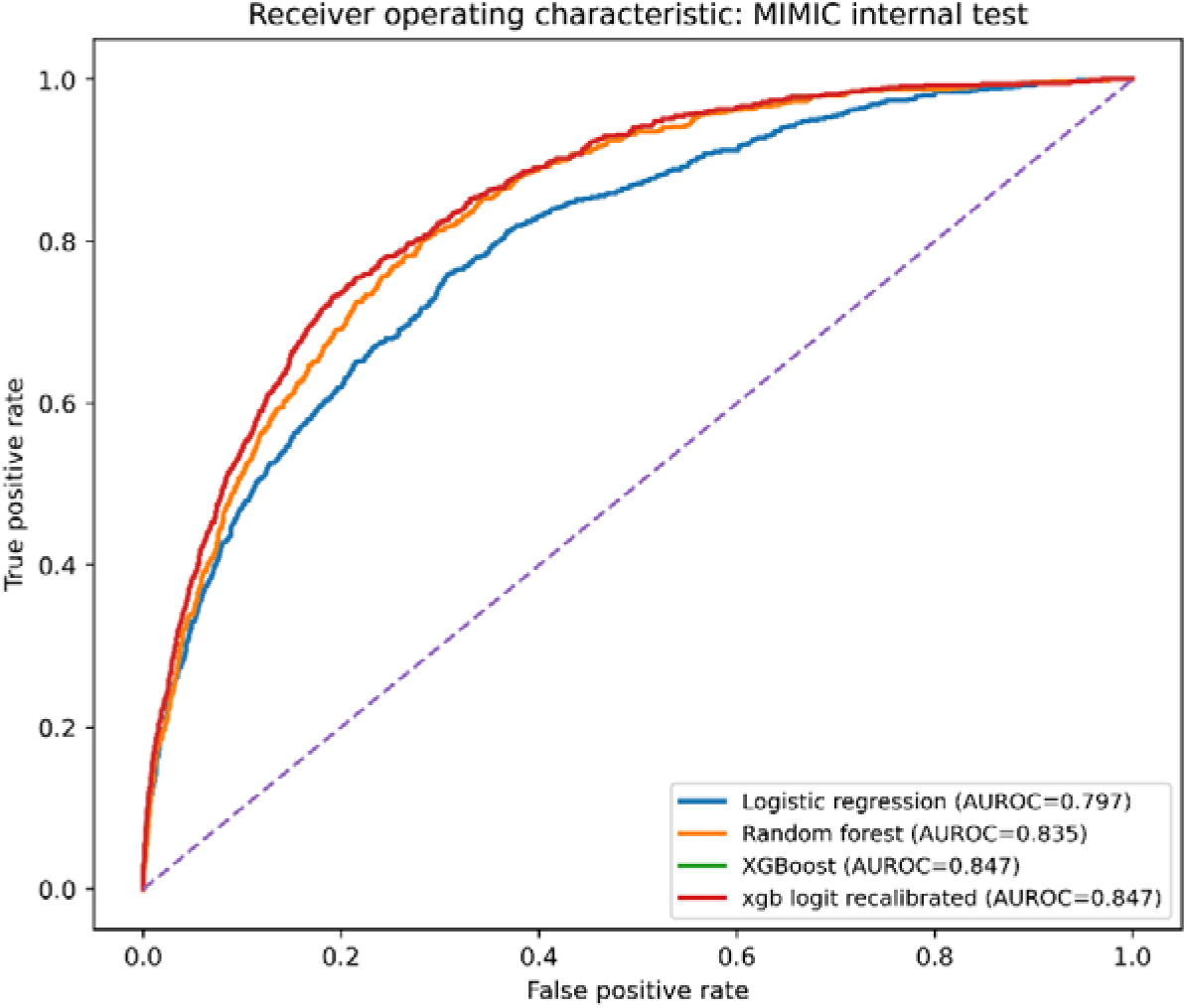
ROC curves for all models on the internal MIMIC-IV held-out test set (n = 7,767). ROC curves for logistic regression (AUROC 0.797), random forest (0.835), XGBoost (0.847), and recalibrated XGBoost (0.847) on the internal test set. The XGBoost and recalibrated XGBoost curves are superimposed because recalibration preserves rank-based discrimination; the two models differ in probability estimates and calibration characteristics.

**Table 2.**
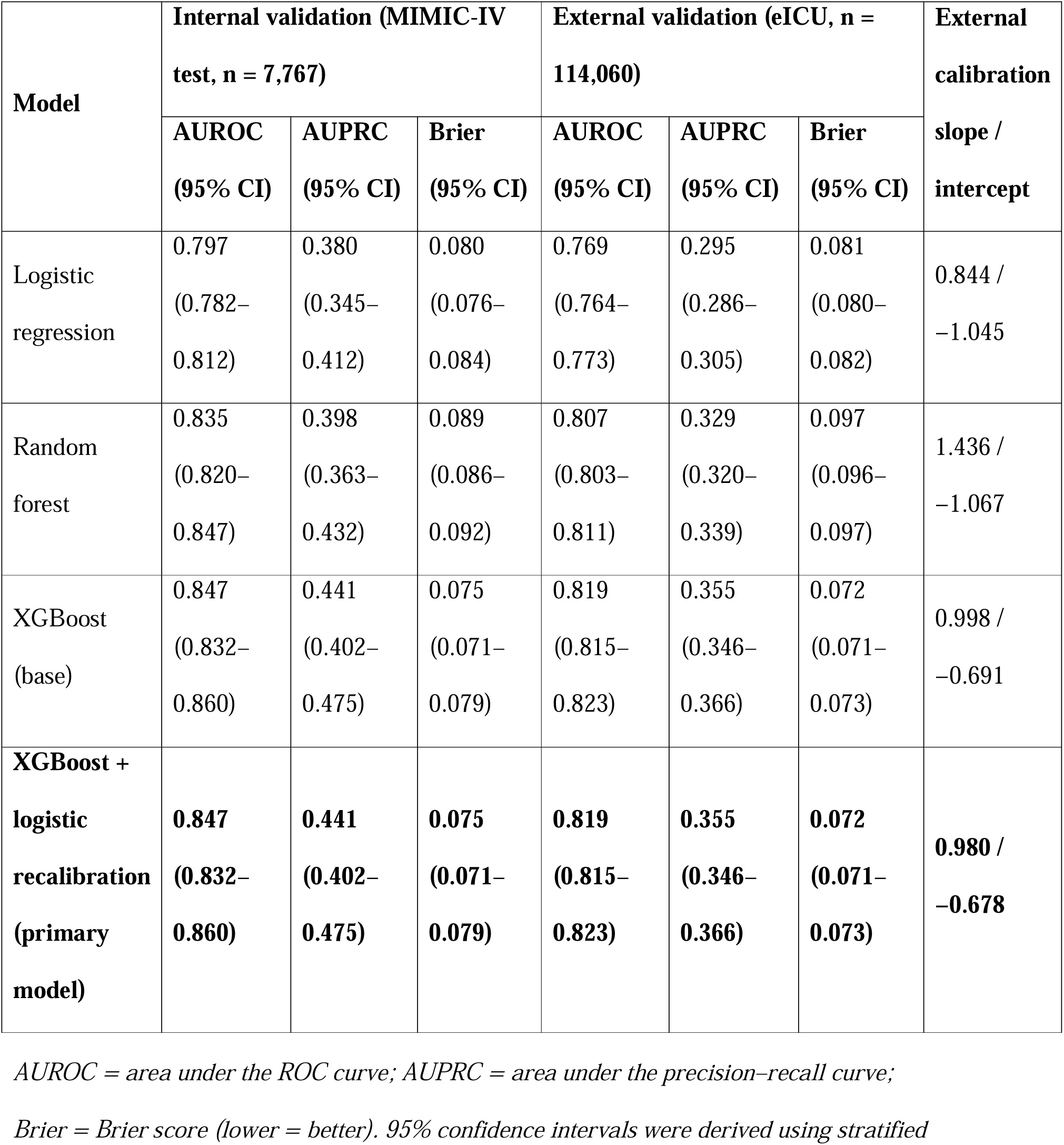
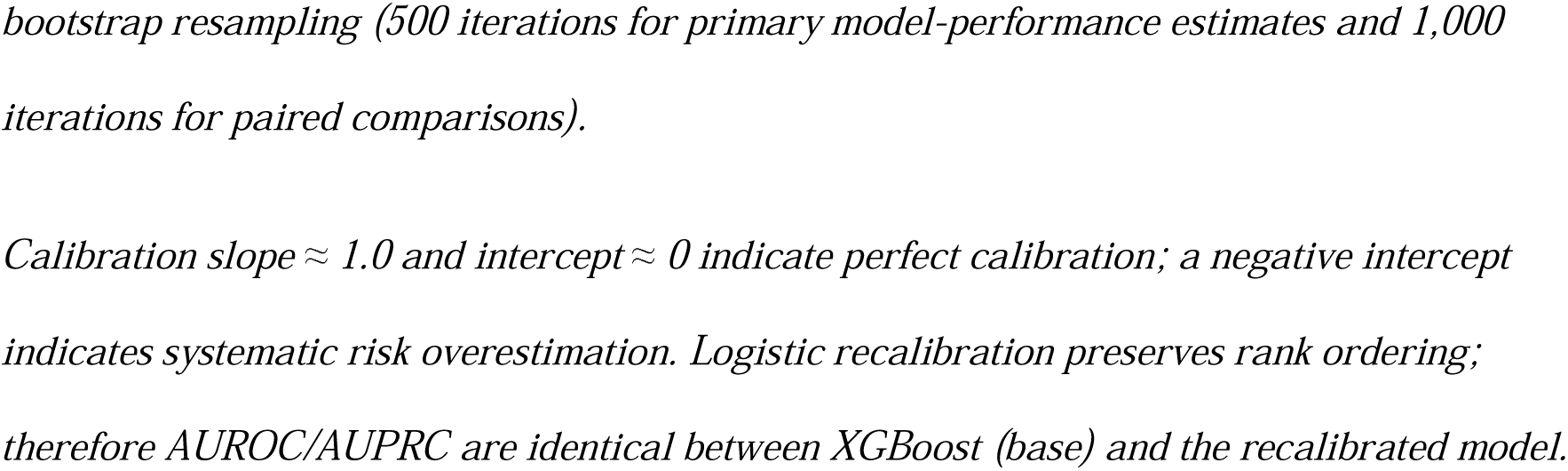
Discrimination, probabilistic accuracy, and external calibration characteristics of all prediction models.

### 3.3 Calibration and Recalibration

Calibration plots are shown in **Figure 3**. Internally, the primary model was near-ideally calibrated (slope 0.982, 95% CI 0.919–1.046; intercept 0.001, 95% CI −0.141 to 0.144; ECE = 0.010). Externally, the calibration slope remained near-ideal (0.980, 95% CI 0.964–0.998), confirming preservation of relative risk ordering across institutions. However, the calibration intercept was substantially negative (−0.678, 95% CI −0.712 to −0.649), indicating systematic overestimation of absolute mortality risk attributable to the 1.81-percentage-point lower event rate in eICU — a pattern consistent with prevalence-driven label shift rather than covariate shift. ECE increased fivefold (0.010 internally to 0.053 externally). A post hoc intercept-only label-shift correction reduced ECE to 0.039 (intercept −0.501), a 26% relative improvement, demonstrating that targeted recalibration without retraining can substantially restore the clinical reliability of probability estimates.

**Figure 3.**
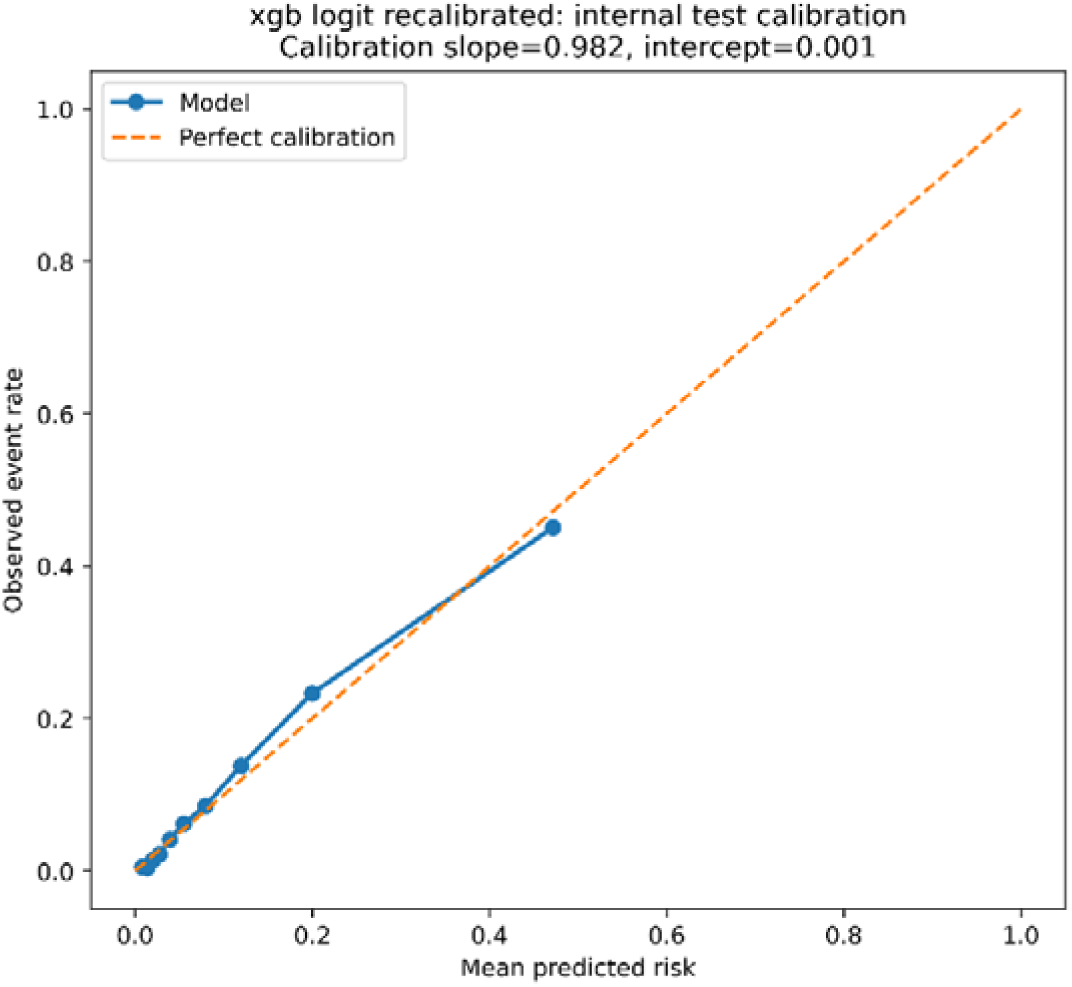
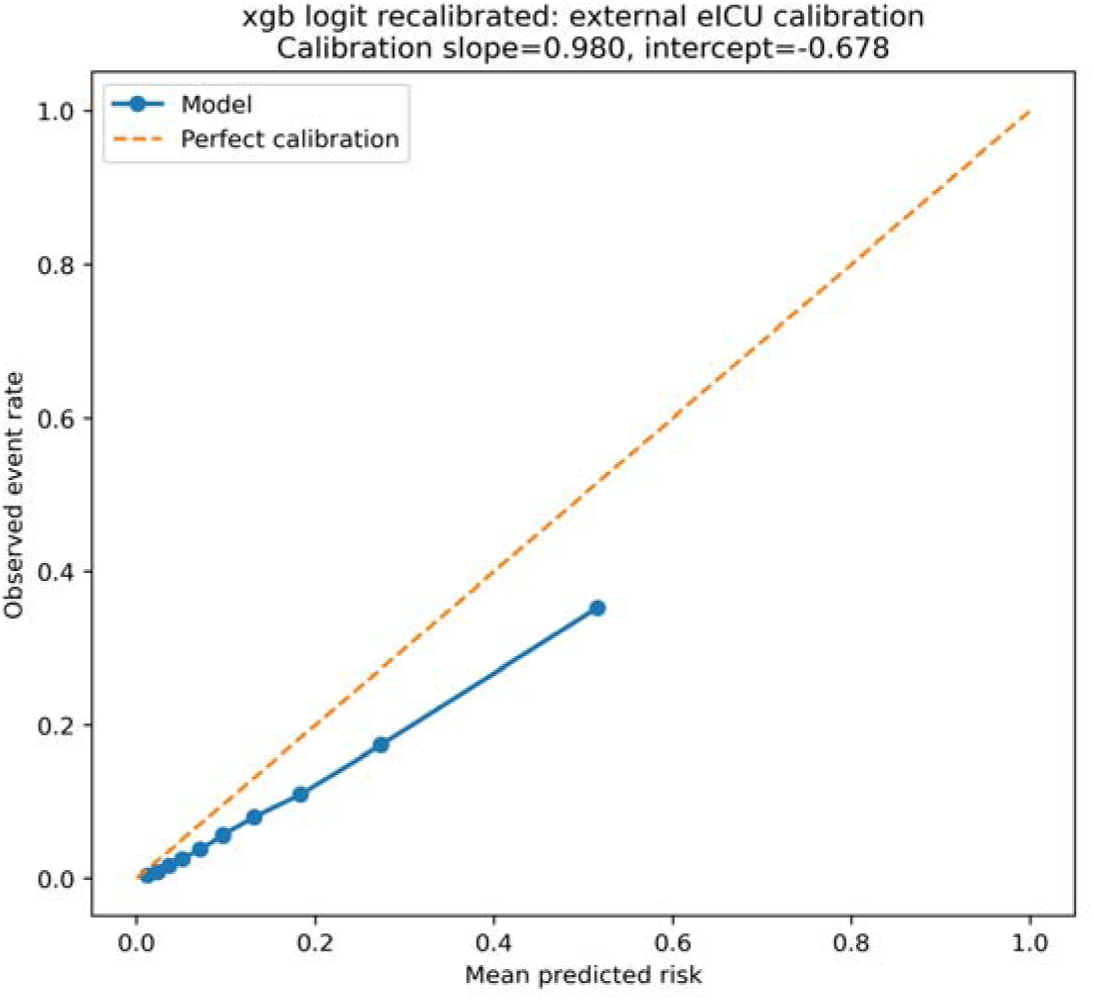
Calibration plots for the primary model on the internal MIMIC-IV test set (upper) and external eICU cohort (lower). Points = mean predicted probability vs observed event rate per decile; dashed orange diagonal = perfect calibration. Internal: slope 0.982, intercept 0.001. External: slope 0.980, intercept −0.678. The near-unit slope externally confirms preserved relative risk ordering; the negative intercept reflects systematic absolute risk overestimation attributable to lower event-rate prevalence in eICU vs MIMIC-IV.

### 3.4 Clinical Utility and APACHE Benchmark

Decision curve analysis demonstrated positive net benefit over treat-all and treat-none strategies across threshold probabilities of approximately 2–40% in the external cohort (**Figure 4**). In the matched eICU subset with available APACHE scores (n = 98,788), the primary model outperformed APACHE in discrimination (AUROC 0.817 vs. 0.795; DeLong p < 0.001) and probabilistic accuracy (Brier 0.074 vs. 0.075). AUPRC was marginally lower (0.364 vs. 0.382), likely reflecting APACHE’s weighting towards high-acuity patients. APACHE exhibited markedly poor absolute calibration (slope 0.591, intercept −1.159), indicating systematic risk overestimation. Full results are in **Table 3**.

**Figure 4.**
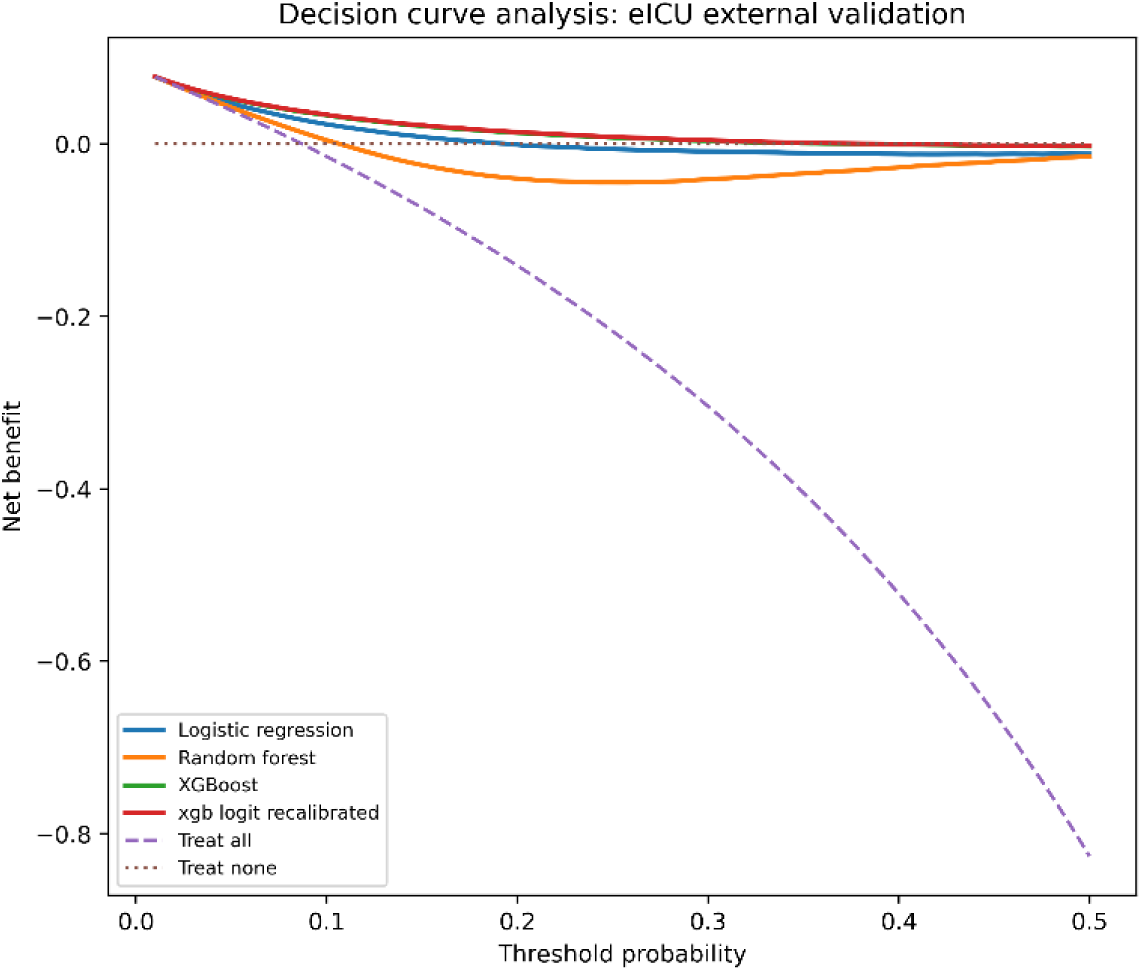
Decision curve analysis in the external eICU cohort (n = 114,060). Net benefit is plotted against threshold probability (0–50%). Dashed purple = treat-all; dotted brown = treat-none. All ML models exceeded treat-none across the full range and exceeded treat-all above ∼5%. The recalibrated XGBoost model yielded the highest net benefit across the clinically relevant 2–40% range. XGBoost and recalibrated XGBoost curves coincide, confirming recalibration does not alter decision-analytic utility.

**Table 3.**
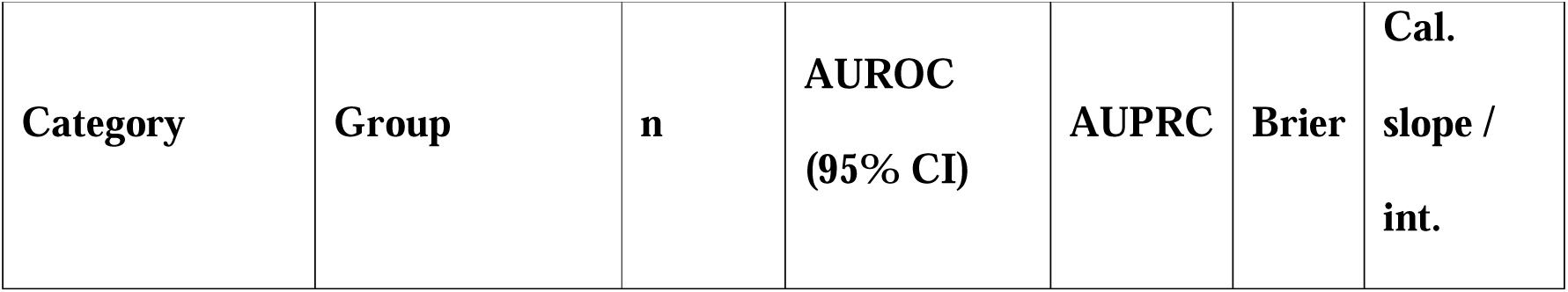
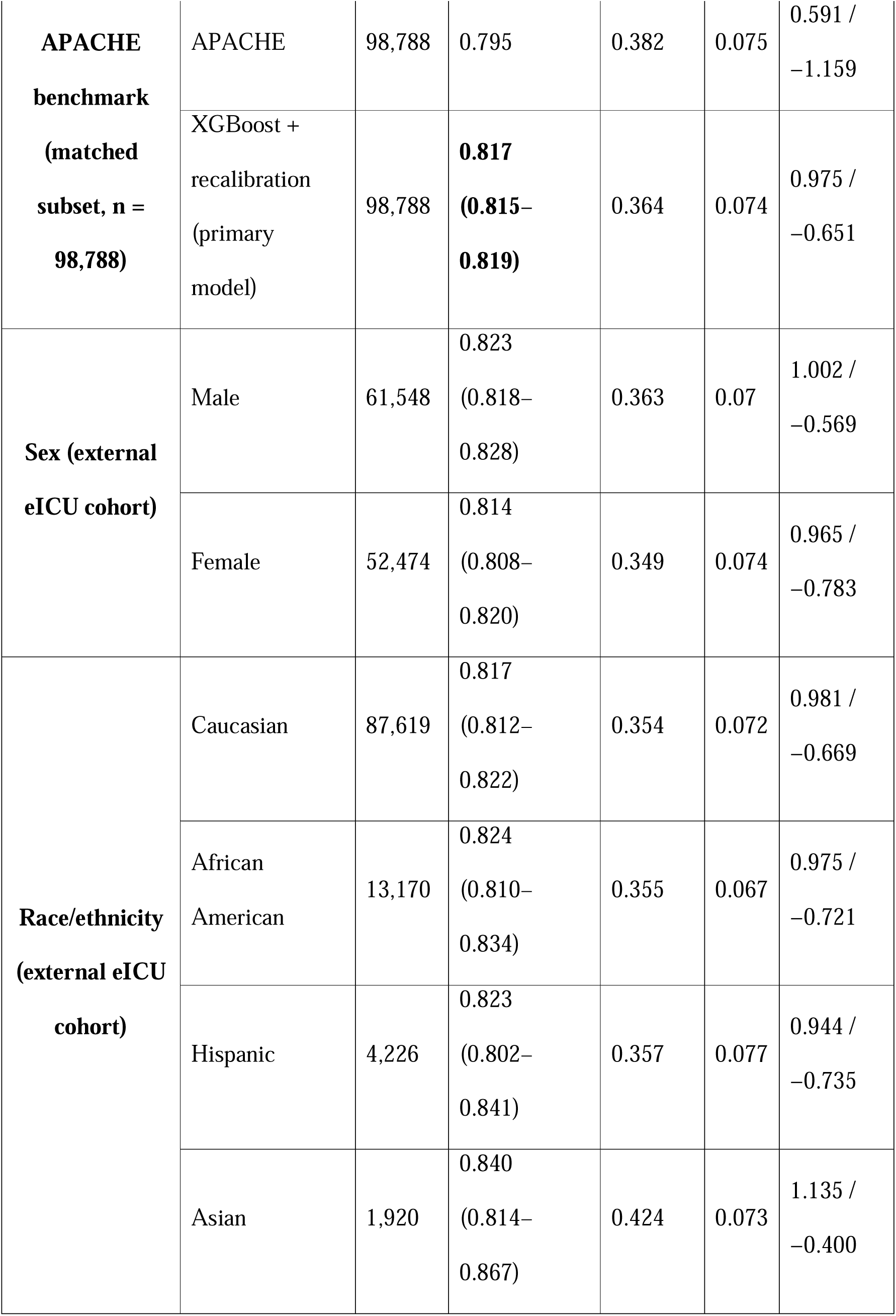
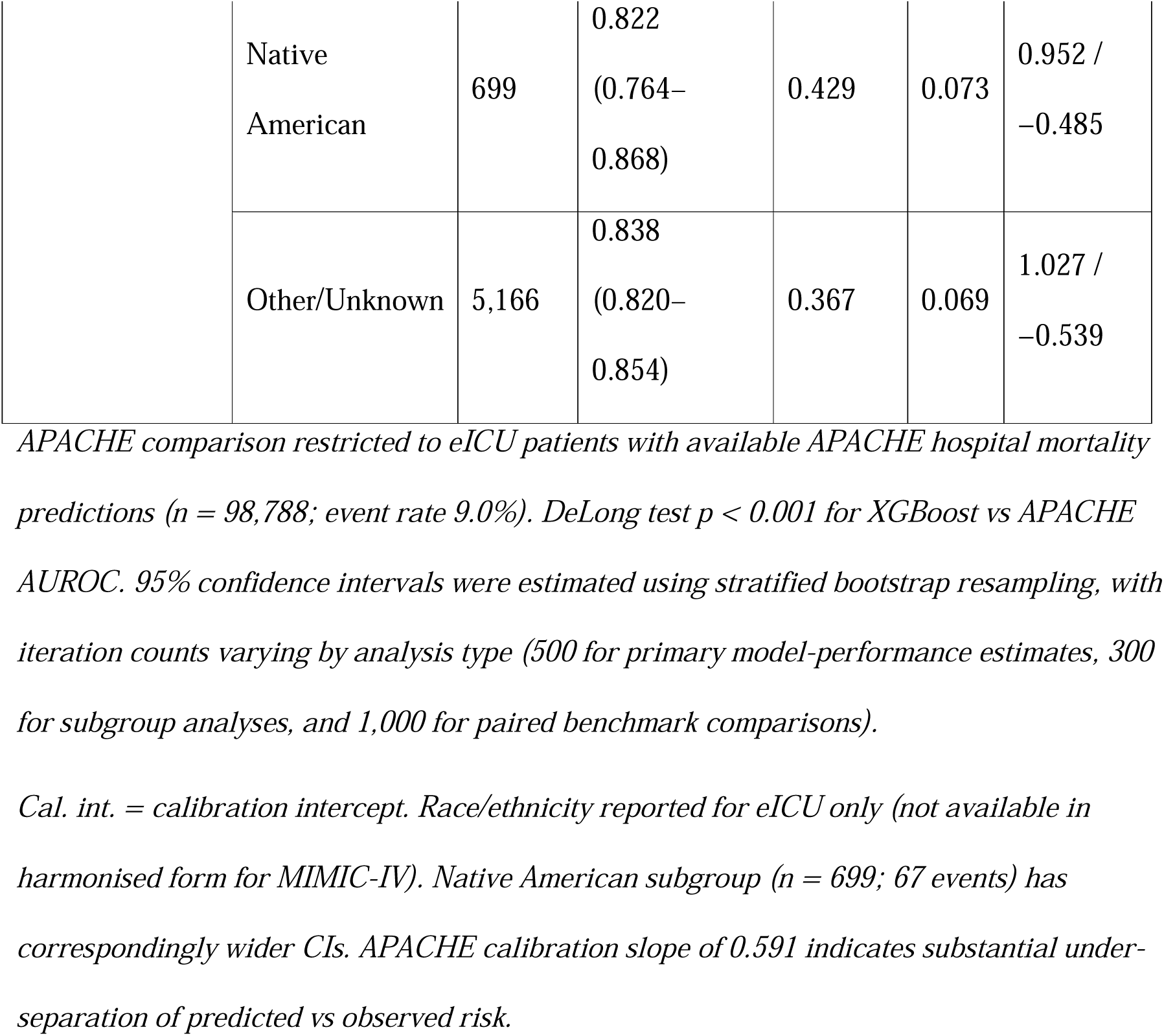
Benchmark comparison with APACHE and subgroup performance of the primary model (external eICU cohort)

### 3.5 Subgroup, Sensitivity, and Secondary Outcomes

Discriminative performance was consistent across sex (AUROC gap 0.009) and racial/ethnic groups (AUROC range 0.817–0.840; maximum gap 0.044), with overlapping confidence intervals for most pairwise comparisons (**Table 3**). Calibration intercepts varied more substantially by subgroup (range −0.400 [Asian] to −0.783 [Female]), indicating unevenly distributed absolute risk overestimation that may require subgroup-specific recalibration prior to deployment. Exclusion of race/ethnicity variables produced negligible change in discrimination (ΔAUROC = +0.001).

Across five sensitivity analyses (Supplementary Table S2), discrimination was broadly stable. A routine-predictor model using only 19 features (excluding arterial blood gas variables) achieved external AUROC 0.794 (ΔAUROC = −0.025), supporting feasibility in resource-limited settings. Restricting to ICU stays ≥48 hours produced the largest attenuation (ΔAUROC = −0.059), consistent with survivor selection bias. For ICU mortality, external AUROC was 0.836 (95% CI 0.830–0.840); for prolonged LOS (≥7 days), external AUROC was 0.720 (95% CI 0.715–0.725).

### 3.6 Model Interpretability

SHAP analysis identified clinically coherent predictors (**Figure 5**). The five highest-ranked features by mean |SHAP| were 24-hour urine output (0.310), age (0.293), maximum BUN (0.291), ventilation flag (0.218), and mean respiratory rate (0.201) — all established markers of organ dysfunction and haemodynamic compromise. Permutation importance yielded consistent rankings (top three: urine output, lactate, age), confirming interpretability robustness. The race variable appeared in the top five by permutation importance (ΔAUROC = 0.0015) but showed modest SHAP contribution, likely reflecting correlation with physiological predictors rather than independent signal. Permutation importance results are in **Supplementary Figure S1**.

**Figure 5.**
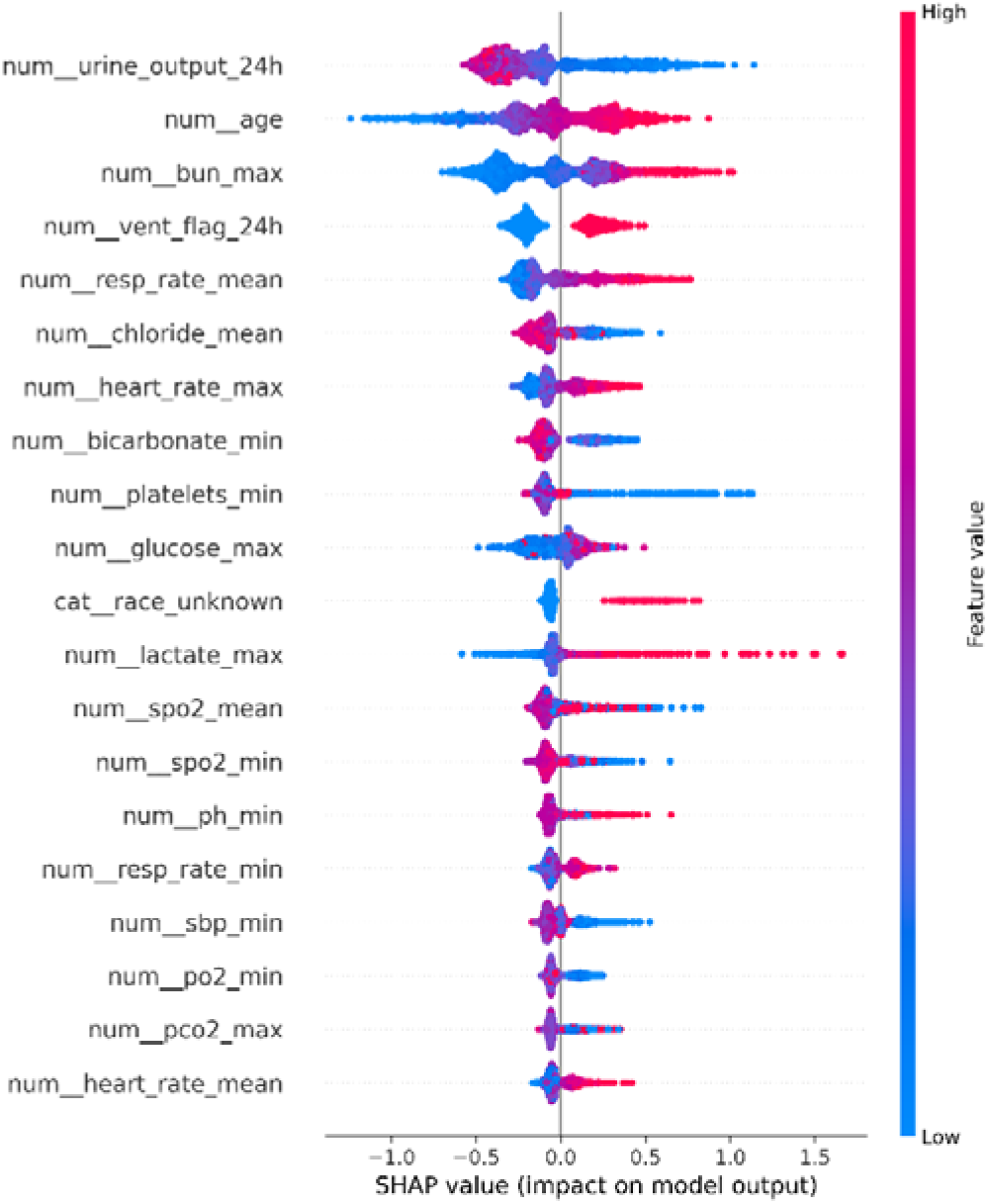
SHAP beeswarm plot for the top 20 features of the primary model (XGBoost + logistic recalibration), computed on a random sample of 2,000 internal test-set observations. Each point represents one ICU stay; x-axis = SHAP value (impact on log-odds of mortality); colour = feature value (red = high, blue = low). Features ranked by descending mean |SHAP|. Feature labels rendered with clinical nomenclature. SHAP values computed using TreeExplainer. Permutation importance is provided in Supplementary Figure S1.

## 4. DISCUSSION

### 4.1 Principal Findings

This study develops and externally validates a reproducible, calibration-aware machine learning framework for ICU hospital mortality prediction and demonstrates that external validation practices relying solely on discrimination metrics may systematically misrepresent model readiness for clinical deployment. Three principal findings emerge.

First, gradient boosting with logistic recalibration achieved transportable discrimination (internal AUROC 0.847, external AUROC 0.819; absolute reduction 0.028), consistent with prior benchmarking studies on MIMIC-derived data reporting AUROC values of 0.82–0.86 using machine learning approaches [3], including deep learning and XGBoost-based methods [8], multitask recurrent architectures [11], and tree-based ensemble models across different critical care outcomes [12]. The present results meaningfully extend prior work through systematic external validation on a fully independent multi-site cohort of 114,060 ICU admissions — validation that remains inconsistently reported across the clinical prediction model literature [13].

Second, the primary model outperformed APACHE in discrimination (AUROC 0.817 vs. 0.795; DeLong p < 0.001) and probabilistic accuracy (Brier 0.074 vs. 0.075) in the matched external subset. APACHE exhibited markedly poor absolute calibration (slope 0.591, intercept −1.159), reflecting a recognized limitation of conventional severity scoring in mixed-acuity ICU populations [3, 13] — reinforcing that well-validated machine learning can offer performance advantages over legacy scores provided calibration is explicitly evaluated prior to deployment [15].

Third, and most critically, discriminative generalizability did not imply calibration generalizability. Despite near-ideal internal calibration (slope 0.982, intercept 0.001), the external calibration intercept was substantially negative (−0.678, 95% CI −0.712 to −0.649), attributable to prevalence-driven label shift — a form of distributional change entirely undetectable by AUROC-based validation. A simple post hoc intercept update, applied without retraining, reduced ECE by 26% (0.053 to 0.039), demonstrating that targeted recalibration can restore clinical reliability at new deployment sites with minimal infrastructure overhead.

### 4.2 Calibration–Discrimination Dissociation and Clinical Utility

The dissociation between preserved discrimination and degraded calibration under cross-institutional shift is mechanistically attributable to the lower in-hospital mortality prevalence in eICU versus MIMIC-IV. When event-rate prevalence differs between development and deployment settings, predicted absolute probabilities diverge from local observed rates even when patient risk rankings are preserved — consistent with emerging evidence that distributional shifts disproportionately affect probability estimates while leaving discriminative ranking relatively intact [15]. Most prior ICU mortality prediction studies report AUROC as the primary or sole metric and do not assess calibration slope and intercept under true external validation [16, 17], leaving a critical gap in deployment readiness assessment [18, 19]. A model with AUROC 0.82 but a calibration intercept of −0.68 may correctly rank the sickest patients while systematically overstating their absolute mortality risk, driving over-intervention near the decision threshold or producing misleading prognostic communications.

Decision curve analysis confirmed positive net benefit across threshold probabilities of approximately 2–40% in the external cohort (Figure 4) [26, 27], encompassing the range most directly relevant to ICU triage, early intervention activation, and resource prioritization. This net benefit, however, is recoverable at new sites only after local intercept recalibration adjusts predicted probabilities to reflect site-specific mortality prevalence. Calibration verification and site-specific recalibration should therefore be treated as prerequisites to deployment rather than optional post-hoc steps. The fully reproducible end-to-end pipeline — encompassing harmonised cohort construction, strict temporal leakage controls, calibration-aware model selection, and public code release — directly addresses the reproducibility concerns identified as structural weaknesses of clinical machine learning research [18, 19].

### 4.3 Equity and Interpretability

Subgroup analyses revealed that while discriminative performance was consistent across racial/ethnic groups (AUROC range 0.817–0.840; maximum gap 0.044) and sex (gap 0.009), calibration intercepts varied substantially by subgroup (range −0.400 [Asian] to −0.783 [Female]) [28, 29]. Absolute risk overestimation was unevenly distributed, indicating that a single global intercept adjustment may not restore equitable probability estimation across all subpopulations. Subgroup-specific calibration monitoring or stratified recalibration protocols should be considered prior to threshold-based deployment [28, 29]. Excluding race/ethnicity variables produced negligible discrimination change (ΔAUROC = +0.001), supporting race-excluded model variants in settings where demographic variables may encode structural inequities rather than independent clinical risk.

SHAP values and permutation importance yielded strongly concordant feature rankings [30], with urine output, age, maximum BUN, ventilation flag, and respiratory rate identified as the five most influential predictors — all established markers of organ dysfunction and haemodynamic compromise. Feature contributions reflect associative rather than causal relationships; clinical interpretation should be made accordingly [30].

### 4.4 Strengths, Limitations, and Future Directions

Key strengths include: two large, diverse public critical care databases (n = 166,088 combined); a comprehensive evaluation framework incorporating discrimination, calibration, decision curve analysis, APACHE benchmarking, fairness analysis, and five pre-specified sensitivity analyses; a fully reproducible pipeline with explicit leakage controls supporting transparent replication [31]; a resource-limited variant (19-predictor model, external AUROC 0.794) demonstrating feasibility in community hospital settings [33]; and public release of all code, model artifacts, and outputs consistent with reproducibility standards for trustworthy clinical machine learning [34]. Decision curve analyses were conducted and reported in accordance with established interpretive guidelines [35].

Limitations include: exclusive reliance on U.S. datasets limiting international generalizability; exclusion of temperature features due to near-complete missingness in MIMIC-IV; use of median imputation rather than multiple imputation; subgroup analyses restricted to sex and race/ethnicity available in eICU; and a retrospective design precluding conclusions about real-world clinical impact. Advanced domain adaptation strategies — including transfer learning and Bayesian updating — were not evaluated. Future work should prioritize prospective stepped-wedge validation within active clinical decision support systems, adaptive site-specific recalibration protocols, and evaluation in non-U.S. and lower-resource healthcare settings to characterize global transportability and broaden fairness assessment.

## 5. CONCLUSION

Machine learning models for ICU mortality prediction can achieve transportable discrimination, but this study demonstrates that transportable discrimination does not guarantee transportable clinical utility. The recalibrated XGBoost model maintained strong external discrimination (AUROC 0.819, 95% CI 0.815–0.823) across 114,060 independent ICU admissions and outperformed APACHE in discrimination and probabilistic accuracy. Yet despite near-ideal internal calibration (slope 0.982, intercept 0.001), the external calibration intercept shifted substantially (−0.678), reflecting systematic absolute risk overestimation driven by prevalence-driven label shift — a dissociation entirely undetectable by AUROC alone. A simple post hoc intercept-only adjustment reduced expected calibration error by 26% without retraining, establishing that deployment-ready calibration is achievable through pragmatically feasible recalibration strategies with direct relevance for health systems deploying predictive tools across institutional boundaries [15, 16].

Calibration must therefore be treated as a mandatory evaluation standard, not an optional reporting item [18, 19]. Equity analyses further demonstrate that calibration intercepts varied substantially across demographic subgroups (range −0.400 to −0.783) despite consistent discrimination, indicating that global recalibration alone may not restore equitable probability estimation — reinforcing the need for subgroup-specific calibration monitoring as a prerequisite to equitable deployment [28, 29]. The reproducible benchmarking framework presented here, developed in accordance with TRIPOD and PROBAST standards [31, 32], a 19-feature resource-limited variant achieving external AUROC 0.794 [33], publicly released code and model artifacts [34], and DCA reporting per established guidelines [35], together provide an immediately adoptable evaluation template for clinical prediction model research in critical care informatics.

Realizing the clinical promise of AI-assisted critical care requires validation that extends unconditionally beyond discrimination to encompass calibration, equity, and decision-analytic utility. Future work should prioritize prospective validation within clinical decision support systems, adaptive site-specific recalibration protocols, and evaluation in non-U.S. and lower-resource healthcare settings to characterize global transportability and prevent amplification of existing disparities in critical care outcomes.

## Supporting information

Supplementary_Table_S1_Sensitivity_Analyses

Supplementary_Table_S2_Secondary_Outcomes

Supplementary_Table_S3_Missingness_Report

Supplementary_Table_S4_Hyperparameter_Tuning

Supplementary_Table_S5_Feature_Selection

Supplementary_Table_S6_TRIPOD_PROBAST_Checklist

## STATEMENTS AND DECLARATIONS

## Data Availability

The datasets analyzed in this study are publicly available. The MIMIC-IV (version 2.2) database and the eICU Collaborative Research Database can be accessed via PhysioNet (https://physionet.org/), subject to completion of the required credentialing, training, and data use agreements. Due to data use restrictions, the datasets cannot be redistributed by the authors. All code and analytical procedures used in this study are publicly available at: https://github.com/Krutarth007/icu-mortality-prediction-ml

## Acknowledgements

The authors would like to acknowledge the contributors of the MIMIC-IV (version 2.2) and the eICU Collaborative Research Database for making these valuable datasets publicly available for research. We also acknowledge PhysioNet for providing access to these resources and supporting reproducible research in critical care.

## Ethical Considerations

This study utilized publicly available, de-identified datasets, namely the MIMIC-IV (version 2.2) and the eICU Collaborative Research Database. Both datasets have received prior institutional review board (IRB) approval, and all patient data were fully de-identified in accordance with the Health Insurance Portability and Accountability Act (HIPAA) Safe Harbor provisions.

As the data are anonymized and publicly accessible, this study was exempt from additional ethical review and did not require informed consent. Access to the datasets was obtained following completion of the required data use agreements and credentialing procedures. All analyses were conducted in accordance with relevant data use policies and ethical guidelines.

## Consent to Participate

Not applicable

## Consent for Publication

Not applicable

## Clinical trial number

Not Applicable

## Use of Generative AI

During the preparation of this manuscript, the authors used generative AI tools (Gemini, ChatGPT, Claude) to assist with language refinement and code debugging. The authors critically reviewed and edited all outputs and take full responsibility for the accuracy and integrity of the final work.

## Author Contributions (CRediT Taxonomy)

- **Krutarth Patel:** Conceptualization (Lead); Methodology (Lead); Software (Lead); Formal Analysis (Lead); Investigation (Lead); Writing – Review & Editing (Lead); Project Administration (Lead).
- **Phanindra Beedala:** Validation (Lead); Data Curation (Lead); Writing – Original Draft (Lead); Methodology (Supporting); Software (Supporting).

---

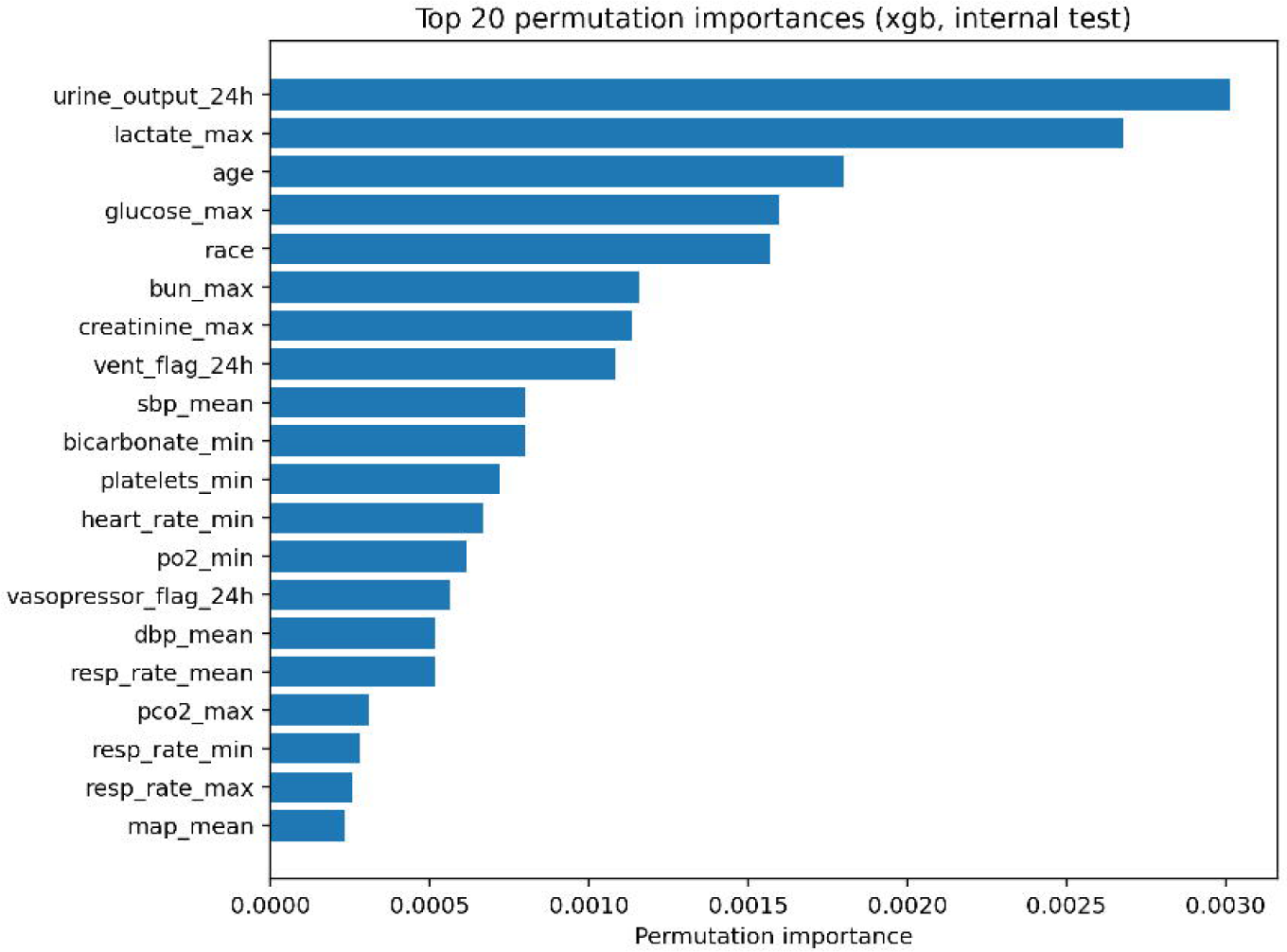

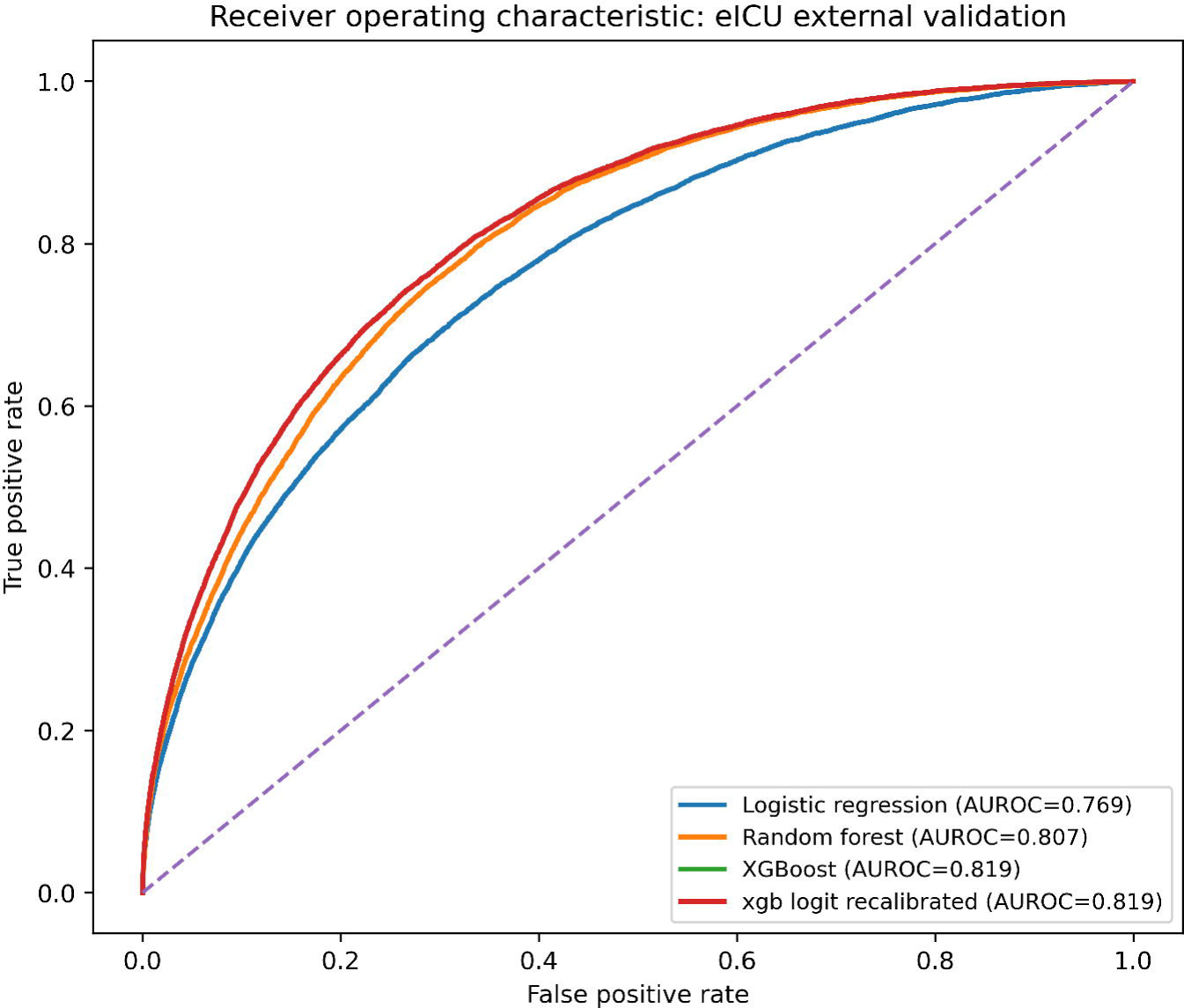

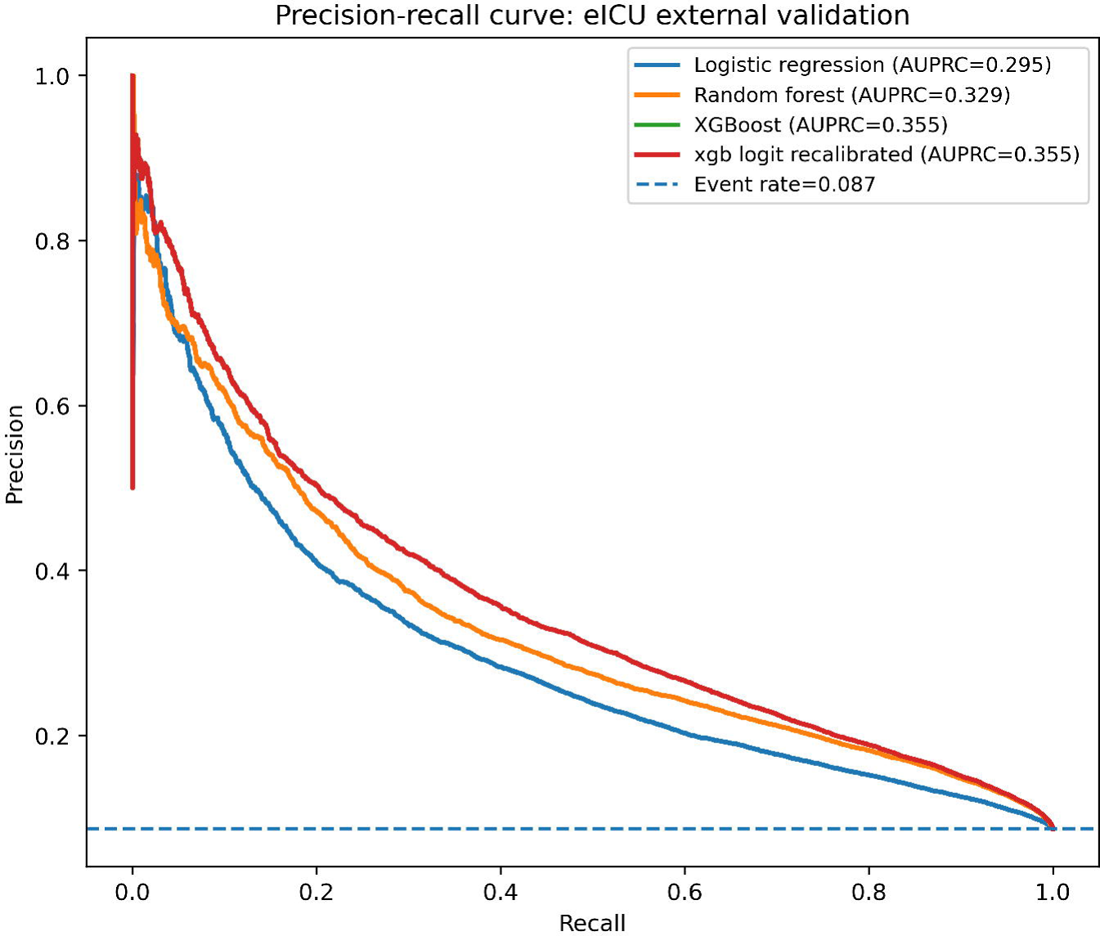

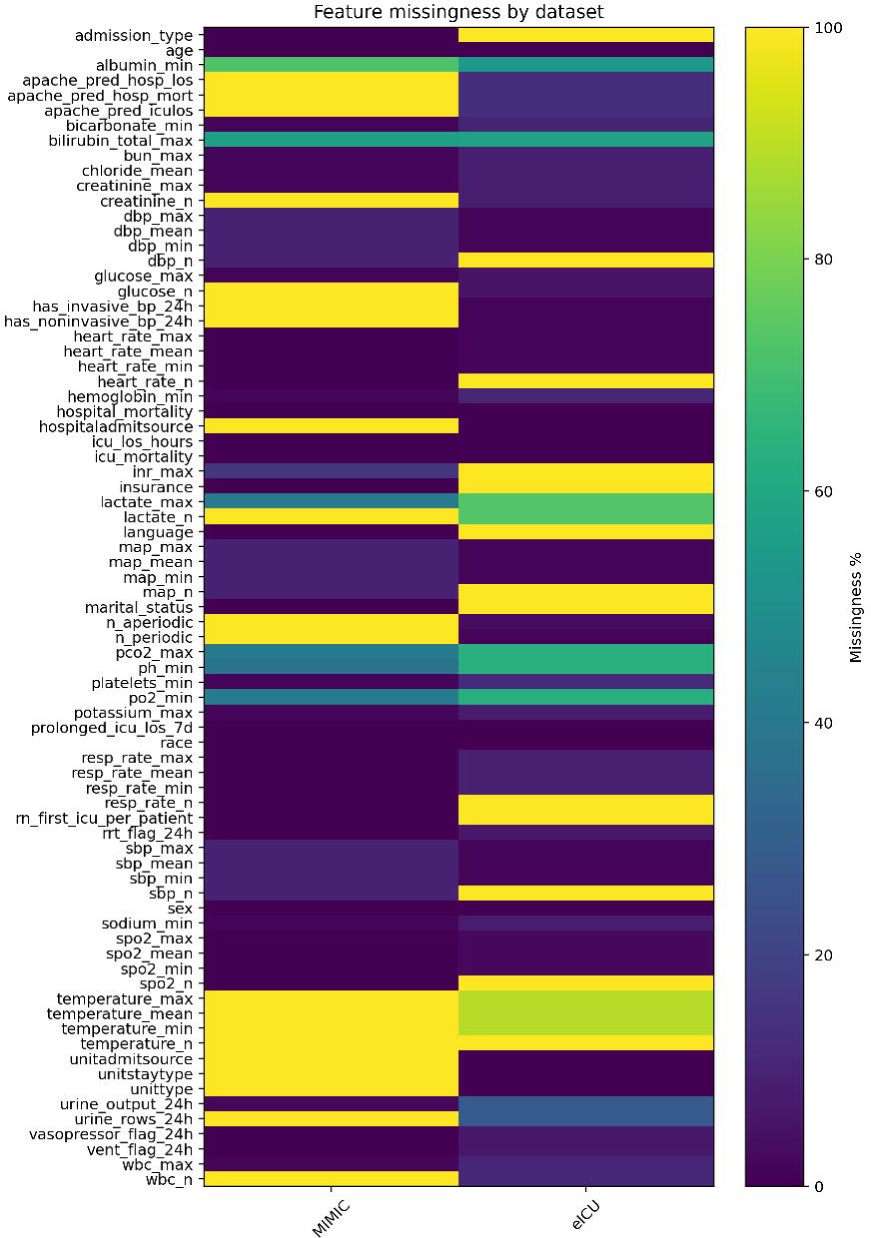

## REFERENCES

1. Komorowski M, Celi LA, Badawi O, Gordon AC, Faisal AA. The artificial intelligence clinician learns optimal treatment strategies for sepsis in intensive care. Nat Med. 2018;24(11):1716–1720. 10.1038/s41591-018-0213-5

2. Shickel B, Tighe PJ, Bihorac A, Rashidi P. Deep EHR: a survey of recent advances in deep learning techniques for electronic health record analysis. J Biomed Inform. 2017;83:168–185. 10.1016/j.jbi.2017.04.001

3. Calvert J, Mao Q, Hoffman JL, Jay M, Desautels T, Mohamadlou H, et al. Using electronic health record collected clinical variables to predict medical intensive care unit mortality. Crit Care Med. 2016;44(2):e61–e67. 10.1097/CCM.0000000000001515

4. Rajkomar A, Dean J, Kohane I. Machine learning in medicine. N Engl J Med. 2019;380(14):1347–1358. 10.1056/NEJMra1814259

5. Zhang Z, Ho KM, Hong Y. Machine learning for the prediction of mortality in patients with sepsis: a systematic review. Ann Transl Med. 2019;7(24):832. 10.21037/atm.2019.11.50

6. Johnson AEW, Ghassemi MM, Nemati S, Niehaus KE, Clifton DA, Clifford GD. Machine learning and decision support in critical care. Proc IEEE. 2016;104(2):444–466. 10.1109/JPROC.2015.2501978

7. Topol EJ. High-performance medicine: the convergence of human and artificial intelligence. Nat Med. 2019;25:44–56. 10.1038/s41591-018-0300-7

8. Purushotham S, Meng C, Che Z, Liu Y. Benchmarking deep learning models on large healthcare datasets. J Biomed Inform. 2018;83:112–134. 10.1016/j.jbi.2018.04.007

9. Johnson AEW, Pollard TJ, Shen L, Lehman LH, Feng M, Ghassemi M, et al. MIMIC-IV, a freely accessible electronic health record dataset. Sci Data. 2023;10:1. 10.1038/s41597-022-01899-x

10. Pollard TJ, Johnson AEW, Raffa JD, Celi LA, Mark RG, Badawi O. The eICU Collaborative Research Database, a freely available multi-center database for critical care research. Sci Data. 2018;5:180178. 10.1038/sdata.2018.178

11. Harutyunyan H, Khachatrian H, Kale DC, Ver Steeg G, Galstyan A. Multitask learning and benchmarking with clinical time series data. In: Advances in Neural Information Processing Systems 32 (NeurIPS 2019); 2019 Dec 8–14; Vancouver, BC. Red Hook, NY: Curran Associates; 2019. Available from: https://proceedings.neurips.cc/paper_files/paper/2019/hash/4735450b461412351b12c3fef0bac8b0-Abstract.html

12. Desautels T, Calvert J, Hoffman J, Jay M, Kerem Y, Shieh L, et al. Prediction of early unplanned intensive care unit readmission using machine learning. Crit Care Med. 2016;44(4):e270–e278. 10.1097/CCM.0000000000001490

13. Steyerberg EW. Clinical prediction models: a practical approach to development, validation, and updating. 2nd ed. New York: Springer; 2019. 10.1007/978-3-030-16399-0

14. Wynants L, Van Calster B, Collins GS, Riley RD, Heinze G, Schuit E, et al. Prediction models for diagnosis and prognosis of COVID-19: systematic review and critical appraisal. BMJ. 2020;369:m1328. 10.1136/bmj.m1328

15. Subbaswamy A, Saria S. From development to deployment: dataset shift, causality, and shift-stable models in health AI. Biostatistics. 2020;21(2):345–352. 10.1093/biostatistics/kxz041

16. Roberts M, Driggs D, Thorpe M, Gilbey J, Yeung M, Ursprung S, et al. Common pitfalls and recommendations for using machine learning in healthcare. Nat Med. 2021;27:745–758. 10.1038/s41591-021-01223-2

17. Kapoor S, Narayanan A. Leakage and the reproducibility crisis in machine-learning-based science. Patterns. 2023;4(8):100804. 10.1016/j.patter.2023.100804

18. Van Calster B, McLernon DJ, Van Smeden M, Wynants L, Steyerberg EW. Calibration: the Achilles heel of predictive analytics. BMC Med. 2019;17(1):230. 10.1186/s12916-019-1466-7

19. Austin PC, Steyerberg EW. The Integrated Calibration Index (ICI) and related metrics for quantifying the calibration of logistic regression models. Stat Med. 2019;38(21):4051–4065. 10.1002/sim.8281

20. Breiman L. Random forests. Mach Learn. 2001;45(1):5–32. 10.1023/A:1010933404324

21. Davis J, Goadrich M. The relationship between precision-recall and ROC curves. In: Proceedings of the 23rd International Conference on Machine Learning (ICML’06); 2006 Jun 25–29; Pittsburgh, PA. New York: ACM; 2006. p. 233–240. 10.1145/1143844.1143874

22. Saito T, Rehmsmeier M. The precision-recall plot is more informative than the ROC plot when evaluating binary classifiers on imbalanced datasets. PLoS One. 2015;10(3):e0118432. 10.1371/journal.pone.0118432

23. Brier GW. Verification of forecasts expressed in terms of probability. Mon Weather Rev. 1950;78(1):1–3. 10.1175/1520-0493(1950)078%3C0001:VOFEIT%3E2.0.CO;2

24. Niculescu-Mizil A, Caruana R. Predicting good probabilities with supervised learning. In: Proceedings of the 22nd International Conference on Machine Learning (ICML’05); 2005 Aug 7–11; Bonn, Germany. New York: ACM; 2005. p. 625–632. 10.1145/1102351.1102430

25. Zadrozny B, Elkan C. Transforming classifier scores into accurate multiclass probability estimates. In: Proceedings of the 8th ACM SIGKDD International Conference on Knowledge Discovery and Data Mining (KDD’02); 2002 Jul 23–26; Edmonton, Alberta, Canada. New York: ACM; 2002. p. 694–699. 10.1145/775047.775151

26. Vickers AJ, Elkin EB. Decision curve analysis: a novel method for evaluating prediction models. Med Decis Making. 2006;26(6):565–574. 10.1177/0272989X06295361

27. Vickers AJ, Van Calster B, Steyerberg EW. Net benefit approaches to the evaluation of prediction models, molecular markers, and diagnostic tests. BMJ. 2016;352:i6. 10.1136/bmj.i6

28. Obermeyer Z, Powers B, Vogeli C, Mullainathan S. Dissecting racial bias in an algorithm used to manage the health of populations. Science. 2019;366(6464):447–453. 10.1126/science.aax2342

29. Chen IY, Szolovits P, Ghassemi M. Can AI help reduce disparities in general medical and mental health care? AMA J Ethics. 2019;21(2):E167–E179. 10.1001/amajethics.2019.167

30. Lundberg SM, Lee SI. A unified approach to interpreting model predictions. In: Advances in Neural Information Processing Systems 30 (NeurIPS 2017); 2017 Dec 4–9; Long Beach, CA. Red Hook, NY: Curran Associates; 2017. Available from: https://proceedings.neurips.cc/paper_files/paper/2017/hash/8a20a8621978632d76c43dfd28b67767-Abstract.html

31. Collins GS, Reitsma JB, Altman DG, Moons KGM. Transparent reporting of a multivariable prediction model for individual prognosis or diagnosis (TRIPOD): the TRIPOD statement. Ann Intern Med. 2015;162(1):55–63. 10.7326/M14-0697

32. Wolff RF, Moons KGM, Riley RD, Whiting PF, Westwood M, Collins GS, et al. PROBAST: a tool to assess the risk of bias and applicability of prediction model studies. Ann Intern Med. 2019;170(1):51–58. 10.7326/M18-1376

33. Kelly CJ, Karthikesalingam A, Suleyman M, Corrado G, King D. Key challenges for delivering clinical impact with artificial intelligence. Lancet Digit Health. 2019;1(6):e312–e315. 10.1016/S2589-7500(19)30123-2

34. Pineau J, Vincent-Lamarre P, Sinha K, Larivière V, Beygelzimer A, d’Alché-Buc F, et al. Improving reproducibility in machine learning research (a report from the NeurIPS 2019 reproducibility program). J Mach Learn Res. 2021;22(164):1–20. Available from: https://jmlr.org/papers/v22/20-1212.html

35. Van Calster B, Wynants L, Verbeek JFM, Verbakel JY, Christodoulou E, Vickers AJ, et al. Reporting and interpreting decision curve analysis: a guide for investigators. BMJ. 2018;362:k3483. 10.1136/bmj.k3483

